# Artifact Identification and Mitigation Strategies for Longitudinal Neural Data Collection Onboard the Medtronic Percept DBS Device^*^

**DOI:** 10.1101/2025.07.23.25331987

**Authors:** Rick R. Hanish, Thomas P. Kutcher, Tomasz M. Frączek, Raphael A. Bechtold, Jeffrey Zhou, Saipravallika Chamarthi, Gabriel Reyes, Mark Libowitz, Ben Shofty, Brian J. Mickey, Brent M. Kious, Wayne K. Goodman, Ankit B. Patel, Sameer A. Sheth, Jeffrey A. Herron, Nicole R. Provenza

**Affiliations:** Electrical and Computer Engineering, Rice University, Houston, TX, USA; Neuroengineering Initiative, Rice University, Houston, TX, USA; Neurosurgery, Baylor College of Medicine, Houston, TX, USA; Bioengineering, University of Washington, Seattle, WA 98195, USA; Bioengineering, Rice University, Houston, TX, USA; Neurosurgery, University of Utah, Salt Lake City, UT, USA; Psychiatry, University of Utah, Salt Lake City, UT, USA; Menninger Department of Psychiatry and Behavioral Sciences, Baylor College of Medicine, Houston, TX, USA; Neuroscience, Baylor College of Medicine, Houston, TX, USA; Neurological Surgery, University of Washington, Seattle, WA, USA

## Abstract

Recent advances in deep brain stimulation (DBS) devices have enabled the ability to capture continuous neural recordings concurrently with stimulation therapy in the background of everyday life. These recordings provide the opportunity to investigate neural biomarkers of various behaviors or clinical status. However, they are susceptible to artifacts that can obscure and limit our ability to interpret neural signals. In a cohort of 23 patients who underwent DBS for obsessive-compulsive disorder (OCD) with the Medtronic Percept device, we identified an artifact in longitudinal neural recordings that occurs when the detected voltage exceeds the device’s maximum sensing capabilities. When such an event occurs, the device inserts a flag value in the neural power stream. We found that overvoltage events are significantly more common in patients implanted with legacy Medtronic 3387 leads than those with newer Medtronic SenSight leads. We demonstrate a best practice, principled strategy for correcting samples affected by overvoltage events to preserve the ability to analyze the data. Finally, in a subset of patients who wore an Oura Ring concurrently with neural recordings (N=14), we found that overvoltage events were more likely during physical activity, suggesting that movement artifacts may elevate low-frequency power regardless of lead model.

## I. Introduction

Recently available commercial deep brain stimulation (DBS) devices enable continuous neural recordings during everyday activities, providing avenues for the longitudinal study of neural activity in natural environments. The ability to study the human brain in ethologically valid contexts has enabled the discovery of neural biomarkers relevant for various psychiatric and neurological disorders [1]–[4]. However, the interpretation of these data can be challenging for several reasons. The data are collected in the real world outside the context of a well-controlled experiment, where confounding variables are difficult to measure [5]. Further, neural data collected onboard DBS devices are known to be susceptible to various types of artifacts, including those resulting from heart activity [6], stimulation [7], [8], and movement [9]. These artifacts impact our ability to effectively analyze neural data and discover accurate and reliable neural biomarkers.

Neural biomarker discovery in Parkinson’s disease has resulted in the recent U.S. FDA approval of adaptive DBS (aDBS) via the Medtronic Percept device [10]. The Medtronic Percept aDBS system monitors chronic 5 Hz-wide local field potential (LFP) band power recordings and adjusts stimulation when the measured LFP power crosses preset thresholds. While stimulation is delivered in an adaptive fashion, information in the data stream that is inaccurate due to an artifact (e.g., movement, EKG) could cause a stimulation parameter change that is unwarranted, potentially worsening symptoms or side effects of DBS. While Parkinson’s disease is the only indication for which aDBS is approved, biomarker discovery in depression [2], obsessive-compulsive disorder (OCD) [1], and other disorders may eventually lead to aDBS trials for additional indications, underscoring the need to effectively mitigate artifacts.

Neural activity collected onboard DBS devices is increasingly being paired with behavioral data from wearables to provide additional context for analyses of neurobehavioral relationships [11], [12] or, in some cases, inform closed-loop DBS parameter changes [13]. Artifacts related to movement are especially relevant for both neurological and psychiatric disorders, as physical activity level may be indicative of clinical status [14], but are also likely to create large amplitude deflections in low frequency neural power [15]. Without longitudinal and synchronized neural and behavioral recordings, it is difficult to tease apart this relationship.

In this study, we analyze data collected from 23 patients with treatment-resistant OCD who underwent DBS of the ventral capsule/ventral striatum (VC/VS). We discovered high-amplitude outliers in low-frequency LFP power recordings that were not biological in nature and to our knowledge not previously been described. Further, we investigated the impact of physical activity and low-frequency LFP power using a wearable device. Finally, we provide a principled strategy for mitigating these high-amplitude artifacts to preserve the ability to analyze the data.

## II. Methods

### A. Study Design

23 adults diagnosed with severe, treatment-resistant OCD underwent DBS surgery targeting the VC/VS region bilaterally after informed consent. All procedures were approved by the local institutional review board (IRB) at the Baylor College of Medicine (BCM; IRB H-48392, IRB H-56119) and the University of Utah (IRB_00169174). Data from 12 of these patients were previously published [1]. In total, we conducted BrainSense Timeline recordings on 44 leads. The remaining 2 leads were in a stimulation configuration that was not compatible with conducting recordings. Eight patients were implanted with legacy Medtronic 3387 leads, and 15 were implanted with Medtronic SenSight leads. Six of the eight patients with legacy leads received the Percept device during a battery replacement procedure that occurred at an average of 3.6 years (*σ* = 1.1 years) after initial DBS. We captured simultaneous Oura Ring and Percept recordings in 14 patients over an average of 4.2 months (*σ* = 4.6 months), comprising an average of over 16,000 samples per patient (*σ* = 2.07 *×* 10^4^).

### B. Medtronic Percept Device BrainSense Timeline Measurements

The BrainSense Timeline function available on the Percept DBS device monitors LFP power in a 5 Hz band once every 10 minutes. Each power measurement is computed on-device via a series of computations. First, the device measures LFP at 250 Hz [16]. After a 10-minute interval *k*, a sliding window is applied to the signal, dividing it into 60 segments, *i*. The power of the user-defined band (in our case, 8.79*±*2.5 Hz, referred to as 9 Hz moving forward) is calculated for each segment, resulting in a set *P*_*k*_ = {*P*_*k*_*i*} *i* 0, 1, …, 59.} Finally, the average power 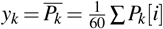 is computed across all segments and stored with the timestamp corresponding to the end of the 10-minute interval.

### C. Overvoltage Events

When the recorded voltage value exceeds the input range of the analog-to-digital converters in the device, the corresponding element in *P*_*k*_[*i*] is filled with the maximum 32-bit unsigned integer value to represent the unknown value. We refer to this occurrence as an “overvoltage event.” Because the maximum integer value is multiple orders of magnitude greater than typical LFP measurements, any overvoltage value drives the average, resulting in:

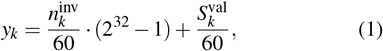

where *y*_*k*_ is the LFP power time series value for interval *k*, 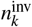 is the number of invalid LFP power measurements *P*_*k*_[*i*] out of 60 that were replaced due to overvoltage events,2^32^ − 1 is the maximum integer value, and 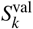 is a relatively small quantity representing the sum of all the valid LFP power recordings (i.e., those that were not overvoltage events) recorded during *k*:

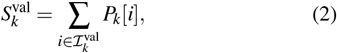

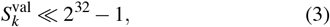

where 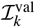 is the set of indices of *P*_*k*_ representing valid (i.e., nonovervoltage) power values. We propose the following three strategies for handling overvoltage events in the data.

#### 1) Thresholding

This overvoltage removal method identifies and removes all values in the stored time series *y*_*k*_ that include at least one overvoltage reading. We identify affected samples using a threshold set at 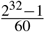.

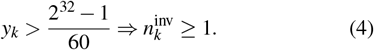

#### 2) Thresholding with Interpolation

Optionally, we may interpolate the missing data with Piecewise Cubic Hermite Interpolating Polynomial (PCHIP) interpolation [17]. This interpolation method uses two valid data points on either side of a segment of consecutive missing data points and their estimated first derivatives to monotonically fill in missing data. When more than 12 consecutive samples (2 hours) were removed or missing, we did not interpolate because simple interpolation is unlikely to capture true neural dynamics over such extended periods.

#### 3) Overvoltage Event Removal (OvER)

Because overvoltage events dominate the average when present, we may estimate how many of the power measurements in *P*_*k*_ were overvoltage events, 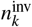, in (5). We then recalculate the interval’s average without the overvoltage events in (6).

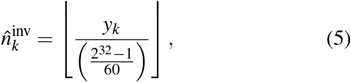

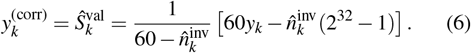

This allows us only to consider valid power recordings, even from within the same 10-minute interval as the overvoltage event, without retaining the overvoltage artifact itself. After using the two replacement methods, we replaced the recorded value in *y*_*k*_ with 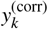 before continuing our analysis.

### D. Data Modality Alignment

To investigate the potential contribution of movement artifacts to neural signals, we leveraged both LFP recordings from the DBS device and movement estimates from the Oura Ring. The Medtronic Percept device provides a continuous LFP power average every 10 minutes. Meanwhile, the Oura Ring estimates a metabolic equivalent (MET) value once per minute. MET represents energy expenditure as a multiple of the patient’s resting metabolic rate (RMR) [18] with a lower bound of 1.0 at rest. Both devices are regularly synced to network time. Given that these two modalities were recorded with different resolutions, we harmonized the data by pairing each neural data point to the maximum MET value over the corresponding 10-minute interval. We chose to use the maximum MET value under the assumption that any movement artifacts in the 10-minute average of low-frequency LFP power are most likely to occur during the instant of highest physical activity.

### E. Statistical Methods

Given that our neural and physical activity data distributions were not normal and exhibited autocorrelation, we conducted significance testing using the Kolmogorov–Smirnov (KS) test with a correction for autocorrelation. We estimated the effective sample size (ESS):

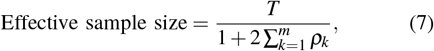

where *T* is the size of the original dataset, and *ρ*_*k*_ represents the autocorrelation coefficient at lag *k*, summed up to a maximum lag *m*. We determined *m* independently for each distribution, selecting the first lag at which its autocorrelation function crossed zero. Each distribution was then downsampled to approximately its estimated ESS to mitigate dependence between samples. Then, we applied the two-sample KS test to the downsampled distributions.

To quantify how strongly physical activity explains increases in neural power, we used Spearman rank correlation to evaluate the relationship between LFP power and associated MET values. The Spearman coefficient is well-suited for our data, which does not meet the normality assumption.

### F. Code Availability

The code to produce the results is available at https://github.com/ProvenzaLab/PerceptArtifactAnalysis.

## III. Results

In 15 of 23 patients, we identified high amplitude deviations in BrainSense Timeline recordings (‘overvoltage events’). Samples affected by overvoltage events appear to form discrete horizontal lines when plotted over time (Fig. 1.a). The number of power measurements that were replaced with the flag value,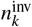, determines the line the measurement is placed on according to (1). Patient data were impacted by overvoltage events to varying degrees. For example, data from patient B009 (left hemisphere) was dominated by overvoltage samples that impacted up to 70% of samples recorded each day (Fig. 1.a). Conversely, data from patient B004 (left hemisphere) had only a handful of overvoltages across their entire duration (Fig. 1.b).

**Fig. 1:**
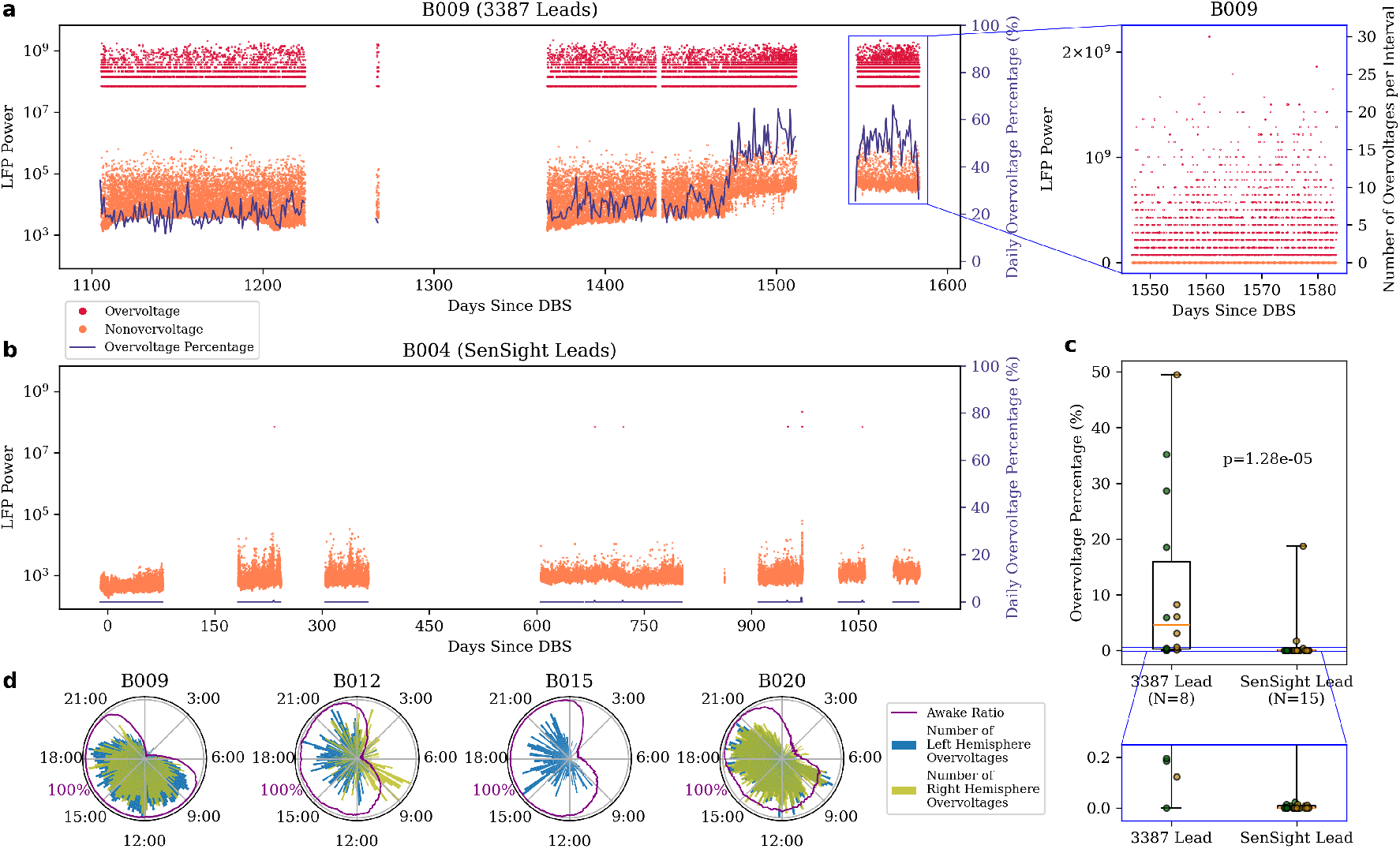
Overvoltage events are primarily observed on Medtronic 3387 leads during waking hours. LFP power time series data recorded from the left hemisphere of patient B009 implanted with 3387 leads (**a**) and B004 implanted with SenSight leads (**b**), overlaid with the daily percentage of samples that include at least one overvoltage event. Up to 70% of recordings per day contain at least one overvoltage for B009 while B004 contains almost none. The right panel of (**a**) shows that overvoltage events form discrete horizontal lines when plotted over time, corresponding to the number of overvoltage events that occurred at each 10-minute interval on the right y-axis. **c**, Box plots show the percentage of samples that contain one or more overvoltage events for each patient with 3387 versus SenSight leads. Green and yellow points correspond to left and right hemisphere leads, respectively. Overvoltage events are more common in patients implanted with 3387 leads than in SenSight leads. **d**, Polar 24-hour plots show overvoltage event counts per 10-minute time bin for each of the four patients with an overvoltage frequency of >5%. The proportion of time patients were awake during each bin derived from Oura ring data is shown by the purple line.

After grouping patients by lead model, we found that overvoltage events were overwhelmingly more common in legacy Medtronic 3387 leads (*µ* = 12.2%) than those with modern SenSight leads (*µ* = 0.50%) (Fig. 1.c).

In patients who experienced an overvoltage in a significant amount (> 5%) of their data, we additionally investigated whether overvoltage events preferentially occurred in the left versus right hemisphere, simultaneously in both, or preferentially during wake versus sleep. Across the dataset, the proportions of samples that contained an overvoltage across the left and right hemispheres were similar (left: 23.4%, right: 11.9%, *p* = 0.78). Further, we found that overvoltages in the left and right hemispheres were not necessarily co-occurring. Only 28.3% of overvoltages in the left hemisphere occurred during the same 10-minute interval as an overvoltage in the right hemisphere. Likewise, only 38.3% of overvoltages in the right hemisphere occurred during the same interval as an overvoltage in the left hemisphere. Lastly, in the four patients with a high overvoltage frequency who provided Oura Ring data, we found that the events occurred preferentially during waking hours (Fig. 1.d).

In testing our mitigation strategies proposed in our methods, the OvER approach was able to best recover the underlying signal. In one example patient, we compare results from correction via OvER (Fig. 2.a) to correction via a simple thresholding and interpolation method (Fig. 2.b). After correcting via OvER, the resulting corrected values appear to preserve a 24-hour neural circadian rhythm (Fig. 2). However, the interpolation method does not reveal a similar structure in the data. To quantify this observation, we fit a cosinor model to the OvER- and threshold-corrected data with a 24-hour period. The fit was significant in all windows across all patients for both correction methods (*p* < 0.020). Critically, we found that the peak cosinor amplitude after correction with OvER was significantly greater than after correction with thresholding and interpolation (*p* < 2.1 × 10^−7^) via a paired *t*-test.

**Fig. 2:**
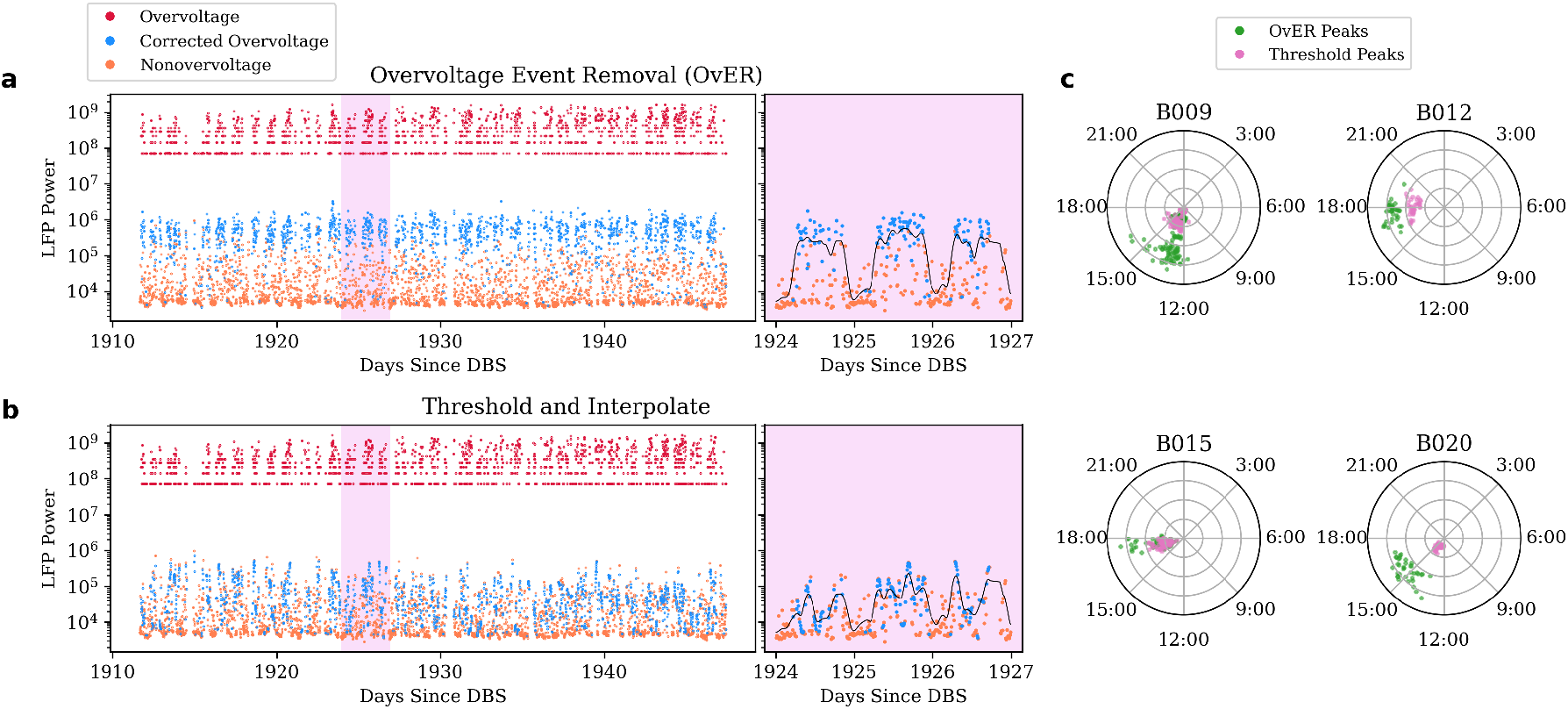
OvER data correction strategy preserves circadian neural patterns. **a**, B020 left hemisphere raw LFP power overvoltages and nonovervoltages are plotted in red and orange, respectively. Corrected overvoltages using the OvER method are overlaid in blue. **b**, Format identical to panel (**a**) with data correction performed using the simple thresholding and interpolation method. The pure thresholding method was not shown here because it simply deletes all overvoltage data without replacement. The right panels of (**a**) and (**b**) show an inset of three days of data along with a Gaussian-smoothed trace to highlight the circadian pattern retained by OvER but not by the interpolation method. **c**, A cosinor model with a 24-hour period was fit to a sliding five-day window of overvoltage-corrected data with a stride of one day. The time of day at which the model’s peak occurred was plotted for each of the four patients with an overvoltage frequency of >5%. This procedure was repeated for both correction methods.

To investigate the relationship between physical activity and overvoltage events, we compared the distribution of MET values captured during overvoltage events and nonovervoltage events. We found that overvoltages were associated with significantly higher METs (*p* = 6.08 × 10^−53^) (Fig. 3.a).

**Fig. 3:**
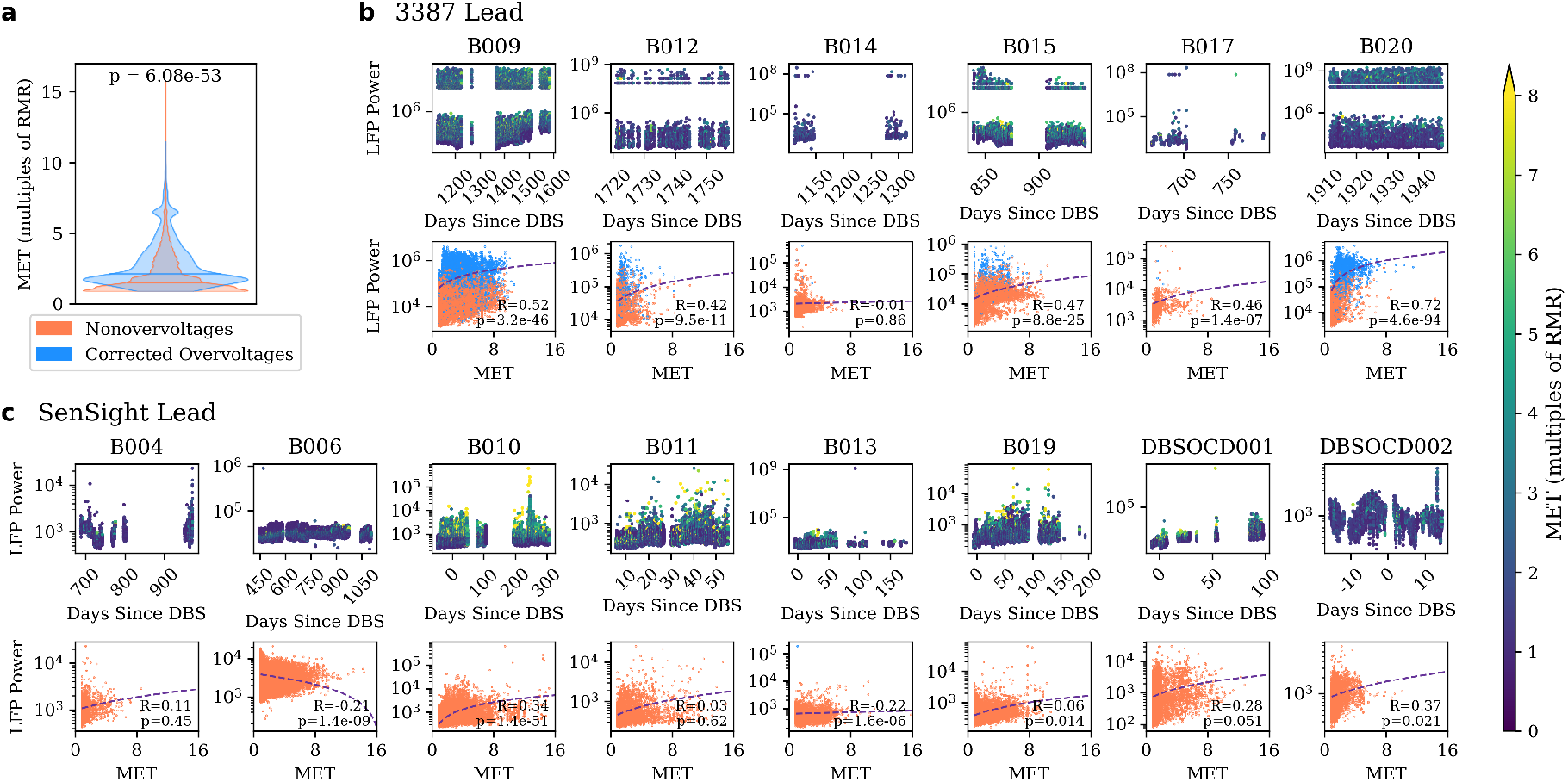
Low-frequency LFP power shows a significant, positive correlation with physical activity in most patients. **a**, The distribution of maximum MET scores that occur during overvoltage events in either hemisphere is significantly greater than the maximum MET scores that occur during nonovervoltage intervals. **b**-**c**, Top plots show the raw time series of left hemisphere LFP power without any correction, colored according to the maximum MET value associated with each sample, per patient. The bottom plots show Spearman correlation between MET and left hemisphere LFP power after correction via the OvER method. Although only left hemisphere data are shown here, right hemisphere results are similar and reported in main text. Panel (**b**) shows patients with 3387 leads, and panel (**c**) shows patients with SenSight leads. Across the entire patient cohort, data from only two leads exhibited a significant, negative correlation between LFP power and MET, six leads had a nonsignificant correlation, and the remaining 17 had a significant, positive correlation.

After adjusting overvoltage values with the OvER method, we explored the relationship between physical activity and neural activity per patient (Fig. 3.b-c). We found a significant, positive correlation between MET and LFP power in 17 of 25 leads (eight left hemisphere, *R* = 0.40, *p* < 0.021), a significant, negative correlation in two leads (both left hemisphere,*R* = −0.22, *p* < 1.6 × 10^−6^), and a nonsignificant correlation in the remaining six leads (four left hemisphere).

## IV. Discussion

We systematically characterized a source of contamination in the 9 Hz LFP power recordings on the Percept DBS device resulting from overvoltage events and provide a best practice strategy for handling these events. We found that overvoltage events disproportionally impact data collected using legacy 3387 versus the newer SenSight lead model. By analyzing Oura Ring MET data collected in parallel with LFP power, we established a small but significant positive correlation between physical activity and low-frequency power, suggesting that physical activity may lead to increased low-frequency power measurements. We also found that overvoltage events are more likely to occur during periods of high physical activity; however, they do not necessarily co-occur in both hemispheres collected on a single DBS device. Together, these findings provide a framework for determining where overvoltages occur in the LFP recorded onboard the Percept device and how to recover the underlying neural signal.

Although new patients are no longer implanted with 3387 leads, they remain commonly used with the Percept device due to how new the SenSight lead model is. One fundamental difference between the data collected via the two lead models is the proportion of overvoltage events collected on each (Fig. 1). Data collected from the 3387 leads had a much greater proportion of overvoltages. While 29 out of 30 SenSight leads were impacted by overvoltage events <5% of the time (with 28 leads impacted <0.1% of the time), one lead exhibited overvoltages at a rate of 18.7%. Further investigation is required to better understand why the overvoltages are occurring so frequently on this single SenSight lead. When collecting BrainSense data on 3387 leads, it is important to quantify the rate of overvoltages occurring in the data and apply a correction strategy.

We propose a strategy for recovering data impacted by overvoltage events that preserves the time series data and enables subsequent time series modeling and feature generation. It exploits the underlying implementation of BrainSense Timeline, in which each reported LFP power value represents an average of 60 individual measurements. The method removes only those measurements flagged as nonneural due to an overvoltage event and re-averages the remaining valid measurements. We found that the OvER method better preserved the neural circadian rhythms that are commonly observed in longitudinal BrainSense Timeline data [1], [19], [20], demonstrating that the artifact correction strategy may significantly impact features estimated downstream. This does not necessarily mean that OvER recovers the underlying signal more accurately than other methods, and each method’s accuracy may not be verified due to the lack of ground truth neural data. Still, the neural circadian rhythms recovered by the OvER method indicate that corrected values retain physiologically relevant fluctuations tied to wake/sleep state, where LFP power is higher during periods of daytime activity and reduced during sleep.

When comparing the distribution of MET values captured by the Oura ring to neural activity recorded by the Percept device, we found that higher levels of physical activity were generally associated with increased low-frequency LFP power. Overvoltage events occurred more frequently during waking hours and tended to coincide with periods of elevated MET. Likewise, a significant positive correlation between MET and LFP power was observed in 17 out of 25 leads. However, this relationship was not consistent across all subjects, with two leads exhibiting a significant negative correlation and six leads showing no significant correlation. Although the positive correlation is weak and not universal, two plausible explanations exist: 1) that increased LFP power during elevated MET is caused by movement artifacts, or 2) that physical activity is accompanied by an increase in 9 Hz power in the VC/VS region. The former explanation is supported by previous work [9], which has shown that neck and jaw movement is related to severe artifacts in recorded LFP data. Additional analysis is required to definitively disambiguate these interpretations.

Future work is required to characterize whether there are specific movements that reliably generate overvoltage events. This could be done via video recordings synchronized to neural activity. While outside the scope of our longitudinal investigations, it is important to investigate how overvoltage events or movement may impact the time domain LFP data stream captured using the BrainSense Streaming modality. Additionally, our future work will investigate the relationship between overvoltage events and stimulation parameters. Some OCD patients decrease DBS amplitude at night, and lower amplitudes may reduce the likelihood of overvoltage events. Lastly, we plan to investigate the consistency of our results in a wider range of frequency bands and target regions beyond the VC/VS. It is possible that the high frequency of overvoltage events that we observed may be unique to our white matter stimulation target and recording regions or the low-frequency band that we are recording. Overall, better characterization and mitigation of overvoltage events will help inform the discovery and refinement of artifact-aware biomarkers in DBS therapy.

## Data Availability

All data produced in the present study are available upon reasonable request to the authors.

